# Diameter measurement of tubular structures on CT angiography, 3D rotational angiography, and 2D digital subtraction angiography against caliper ground truth: A phantom study of surrogates for intracranial vessels

**DOI:** 10.64898/2026.09.05.26362337

**Authors:** MD Alexander, F Settecase

## Abstract

**BACKGROUND AND PURPOSE:** Measured intracranial vessel dimensions guide device selection in cerebrovascular intervention, yet accuracy depends on imaging modality, luminal contrast concentration, and the lumen-edge detection criteria. Prior accuracy studies benchmark one angiographic modality against another rather than physical ground truth. This study quantifies accuracy of CT angiography (CTA), 3D rotational angiography (3DRA), and 2D digital subtraction angiography (DSA) against a phantom of tubular contrast-filled structures serving as surrogates for intracranial vessels.

**MATERIALS AND METHODS:** Phantoms with cylindrical lumen diameters 1.2–8.5 mm at graded iodinated contrast concentrations (6.25–100%) were imaged with clinical devices in air and a tissue-equivalent gel surround. Tubular cross-sections perpendicular to centerlines were measured by maximum-gradient, full-width-half-maximum (FWHM), and fixed-Hounsfield criteria and compared with physical phantom measurements using mixed-effects models.

**RESULTS:** Across 1682 cross-sections (73 lumens), all modalities recovered true diameter within ∼0.5 mm but consistently measured larger than the true dimensions. Over-measurement occurred minimally in the 2–5 mm range, maximally at the extremes of caliber and contrast concentration. 3DRA was modestly closer to physical phantom measurements than CTA (gradient bias +0.40 vs +0.60 mm). Maximum-gradient edge over-measured diameters versus the conventionally preferred FWHM, with the latter carrying larger, more concentration-sensitive bias. Fixed-HU thresholds were concentration-biased on CTA and unusable on 3DRA. 2D DSA, currently the clinical reference standard, was itself biased in analysis of its auto-calibrated measurements, over-measuring large lumens, under-measuring small lumens 1.5 mm, and reading larger frontally than laterally through magnification.

**CONCLUSIONS:** Conventional CTA, 3DRA, and 2D DSA each measure intracranial-scale caliber lumens within ∼0.5 mm but with consistent bias. Maximum-gradient lumen edge detection minimizes bias and outperforms FWHM, while fixed-Hounsfield contour tracing is unreliable, particularly on 3DRA. Characterizing these error modes provides a calibration basis for automated vessel-measurement platforms.

## INTRODUCTION

Accurate intracranial vascular dimensions are important for planning and performing cerebrovascular interventions. For instance, in mechanical thrombectomy, under-sizing an aspiration catheter compared to the inner diameter of the occluded artery can lead futile aspiration attempts, while over-sized devices can cause vascular damage potentially leading to serious complications^1,2^. Flow diversion treatment for intracranial aneurysm treatment is similarly reliant on device sizing for safe and effective treatment. Oversizing reduces occlusion rates due to incomplete expansion and increased porosity^3,4^. More recent work indicates oversizing likely triggers smooth muscle cell hypertrophy that subsequently may contribute to braid deformation on follow up^5^. Undersized flow diverting stents can migrate if substantial size mismatch exists, while non-apposition and the ends of the device can induce flow stasis, platelet aggregation, and thromboembolic complications^6,7^. Similarly, venous measurements require accuracy for treatment device selection, such as for venous sinus stenting for idiopathic intracranial hypertension^8^. Imaging platforms that measure intracranial vessels with the aim of treatment planning must therefore measure vessel dimensions accurately and precisely across a range of calibers for vessels most commonly treated in the intracranial circulation, i.e. ∼1-10 mm.

Measured diameter is a property of the imaging chain that must be handled in ways that most closely represent the ground truth dimensions of a vessel. Partial-volume averaging, especially in computed tomography reconstruction, blurs the transition from contrast in the lumen to adjacent structures, rendering the wall of the vessel itself thin and often imperceptible. This determination of the lumen edge affects the measured diameter, so placement of PACS software calipers at this edge is of utmost importance for correct vessel dimension measurements. Luminal contrast concentration sets the height of that transition from vessel to non-vessel pixels (or voxels); CTA is known to underestimate large arteries and overestimate small vessels relative to angiographic references, with prior intracranial artery studies showing CTA measurements to be approximately 0.26 mm below measurements on 2D DSA^9,10^. When measuring vessel characteristics on cross-sectional short axis views, diameter, area, or circularity will be affected by the lumen-edge detection criteria employed. These criteria, such as fixed Hounsfield threshold, FWHM, or gradient edge detection, determine the contour of a vessel on a given image. Of these lumen edge detection criteria, FWHM is the established partial-volume-corrected criterion^11^, and on 3DRA gradient edge detection outperforms fixed thresholds for aneurysm volumetry^12^.

3DRA is geometrically accurate on phantoms but is susceptible to thresholding and windowing dependency^13,14^. Phantom-based calibration of CT lumen quantification has been performed largely in the coronary circulation^15^. Cerebral CTA–3DRA work has predominantly compared aneurysm detection and geometry rather than analysis of normal parent vessel dimensional features^16,17^. Investigation for intracranial imaging purposes has relied on comparison between imaging modalities rather than comparison against a structure with known physical measurements, a consequence of the inability to measure intracranial vessels in living humans directly. As such, dedicated partial-volume calibration of the lumen-edge criterion for small intracranial vessel diameters across both CTA and 3DRA is, so far, lacking. Nearly all prior accuracy studies use 2D DSA or 3DRA as a reference standard^9,10,16,17^. This circular logic cannot establish which modality is most accurate, nor whether the inter-modality inaccuracy can be anticipated and corrected. The current study compares 2D DSA, 3DRA, and CTA measurements against tubular phantom structures with physically measured dimensions, with the primary aim to establish the accuracy of CTA, 3DRA, and 2D DSA and whether they mis-measure in a structured, predictable way that can assist calibration.

## MATERIALS AND METHODS

Two phantoms of contrast-filled cylindrical lumens of known inner diameter were constructed (Fig 1, Fig 2). Phantom 1 comprised 16 syringes of four types (0.5-mL and 1-mL standard syringes; 3-mL arterial-blood-gas [ABG] and 3-mL Medallion syringes) filled with iodixanol-320 at four concentrations (12.5%, 25%, 50%, 100%) and imaged while sandwiched in horizontal position between two pieces of foam. Phantom 2 comprised 3-mL Medallion and 1-mL standard syringes and small-bore extension tubing (smallest lumen, ∼1.2 mm) at five concentrations (6.25–100%), positioned vertically and arranged radially in a circular plastic basin for support. Phantom 2 was used with two surrounds in addition to the plastic from the basin—air (leaving the basin empty) and a gel substrate into which phantom objects were placed.

**FIG 1.**
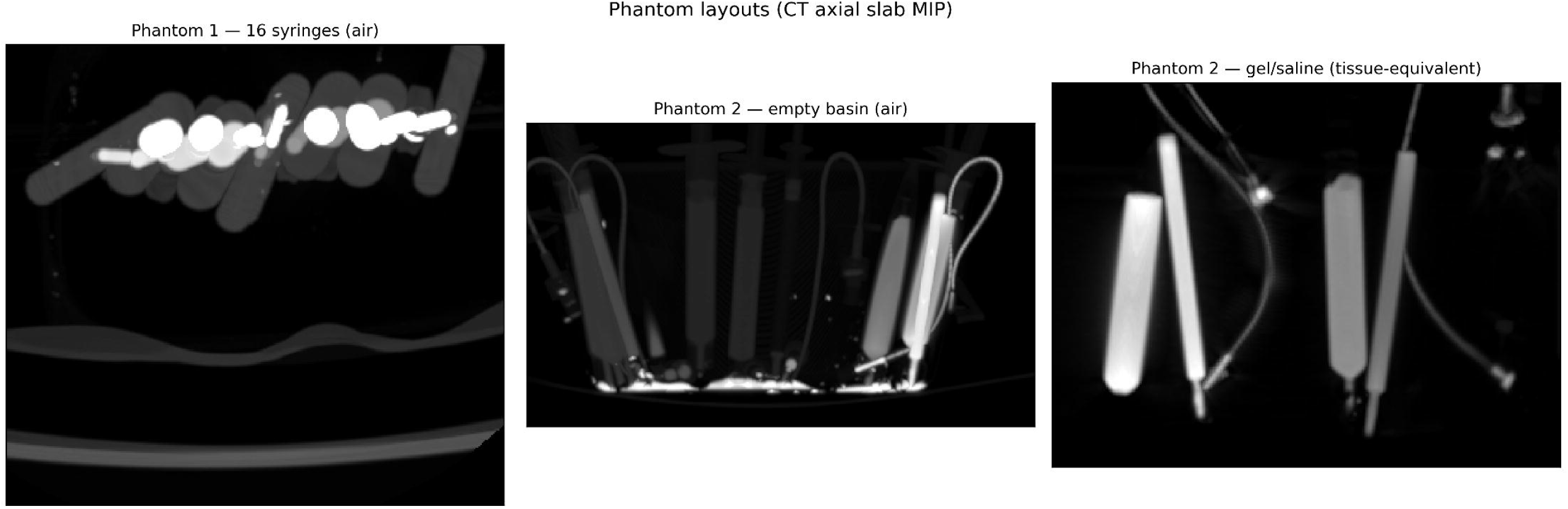
Phantom layouts (CT axial slab maximum-intensity projection): phantom 1 (16 syringes, air); phantom 2 empty basin (air); phantom 2 gel/saline (tissue-equivalent).

**FIG 2.**
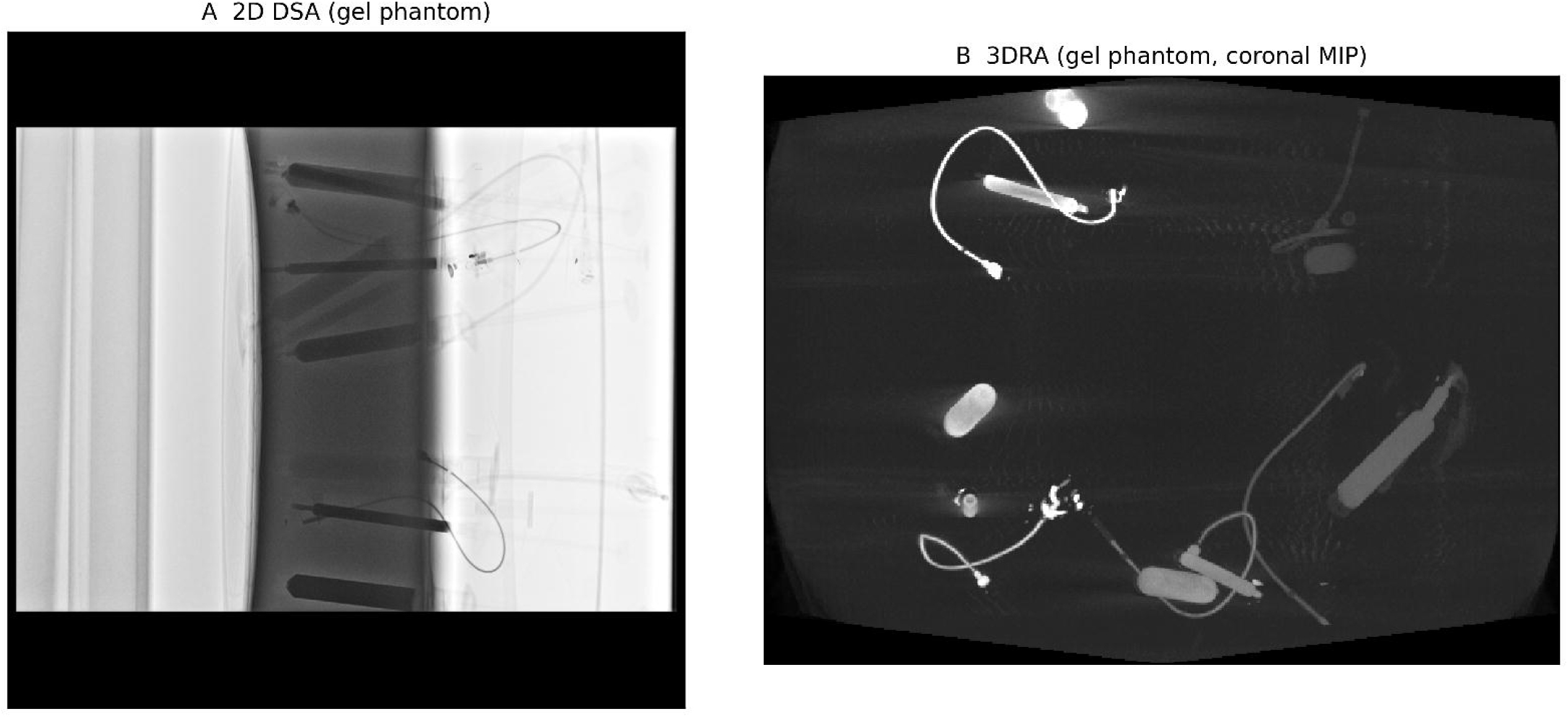
Representative gel-phantom images. A, 2D DSA frontal projection. B, 3DRA coronal maximum-intensity projection of the same tissue-equivalent phantom, showing each modality’s appearance of the contrast-filled syringes and extension tubing.

After imaging, each lumen was sectioned and the inner bore measured with digital calipers (Fig 3) at each of the two cut faces and two orthogonal axes per face (four measurements per lumen; mean ± SD). Cutting was performed with a high-speed rotary tool for the hard plastic syringes and surgical scissors for the extension tubing. Mid-bore (cut-face) measurement matched the mid-body region sampled in the images and avoided the wider plunger-insertion lip. The orthogonal measurements were obtained to account for any deformation that may have occurred from the cutting process or any preexisting non-uniformity of the cylinder, allowing for averaging to of orthogonal dimensions to converge at the diameter of the cylinder to be measured on the imaging modalities.

**FIG 3.**
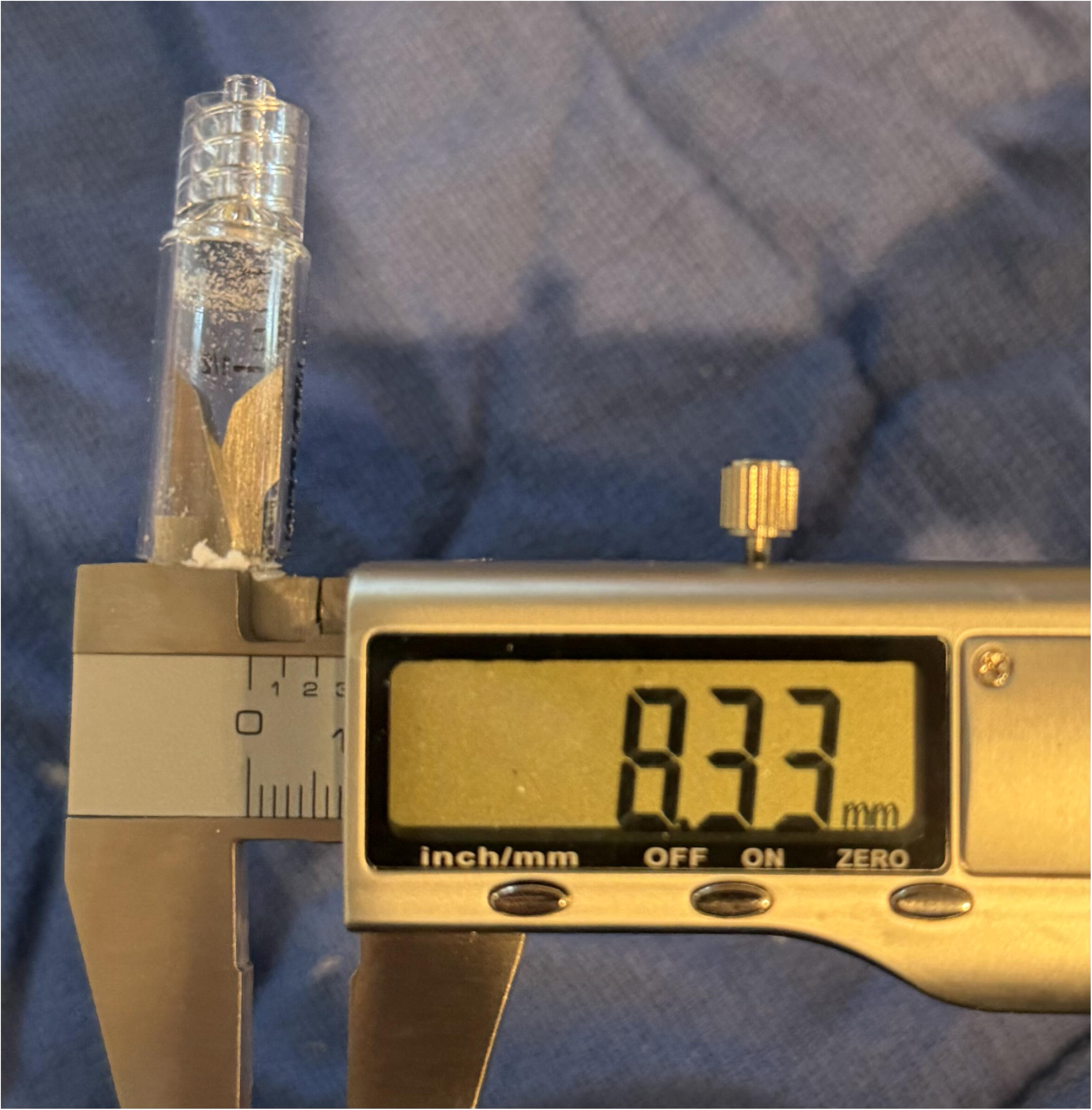
Photograph of digital calipers measuring the cut edge of a plastic syringe.

CT acquisitions were obtained according to clinical algorithms used for patients undergoing evaluation for suspected acute ischemic stroke and scanners currently in use clinically. Phantom 1 was imaged on a GE LightSpeed Ultra with 120 kVp and 0.625 mm slice thickness and a standard kernel. Phantom 2 was imaged on a Siemens SOMATOM X.cite with 120 kVp and 0.5-mm slice thickness and Hr40s kernel. Both phantoms were also imaged on a Siemens AXIOM-Artis ICONO biplane unit, acquiring images for 3DRA using the unit’s 4sDSA Head acquisition and processing the resulting native-fill output with full-HU-auto setting and 0.481-mm isotropic geometry. The phantoms were also imaged with conventional 2D DSA on the biplane unit in frontal (≈0°) and lateral (≈90°) projections. Diameters were measured on these 2D images by reading the magnification-corrected image using each frame’s isocenter-calibrated pixel spacing (detector pitch ÷ source-to-detector/source-to-isocenter magnification, 1.49–1.65×), so residual error would reflect off-isocenter magnification. Representative gel-phantom images on both 2D DSA and 3DRA are shown in Fig 2. Representative gel-phantom images on both 2D DSA and 3DRA are shown in Fig 2.

For the lumen of each device, a centerline was defined from manually placed waypoints (3 in straight syringes and 5-8 in curved tubing), interpolated piecewise-linearly for straight objects and by cubic spline for tubing so that sampling planes remained orthogonal through bends. Cross-sections were sampled perpendicular to the local tangent at 2-mm intervals at 0.10-mm in-plane resolution.

On each cross-section, after recentering on the bright lumen, inner diameter was estimated by (1) a maximum-gradient edge (radius of steepest intensity drop along radial profiles, background-independent); (2) FWHM (radius at 50% between luminal peak and background); and (3) fixed-Hounsfield thresholds (200/300/500 HU on CTA; 800/1500/3000 on 3DRA). Cross-sections were excluded as non-measurable if the diameter exceeded 13 mm. To exclude non-orthogonal cross-sections through curves of tubing, those with eccentricity exceeding 0.5 were excluded.

Cross-sections were also excluded if the luminal peak fell below an absolute brightness threshold, indicating leakage-affected lumens. Additionally, lumens with fewer than five valid sections were excluded. On the 2D DSA projections, where overlapping objects preclude automated cross-section sampling, lumen width was instead measured manually edge-to-edge at three non-overlapping levels per object (145 widths total), with frontal and lateral views giving two orthogonal axes. Only objects clearly separable from neighbors in each projection were measured. The gradient and FWHM contours on representative lumen cross-sections are shown in Fig 4.

**FIG 4.**
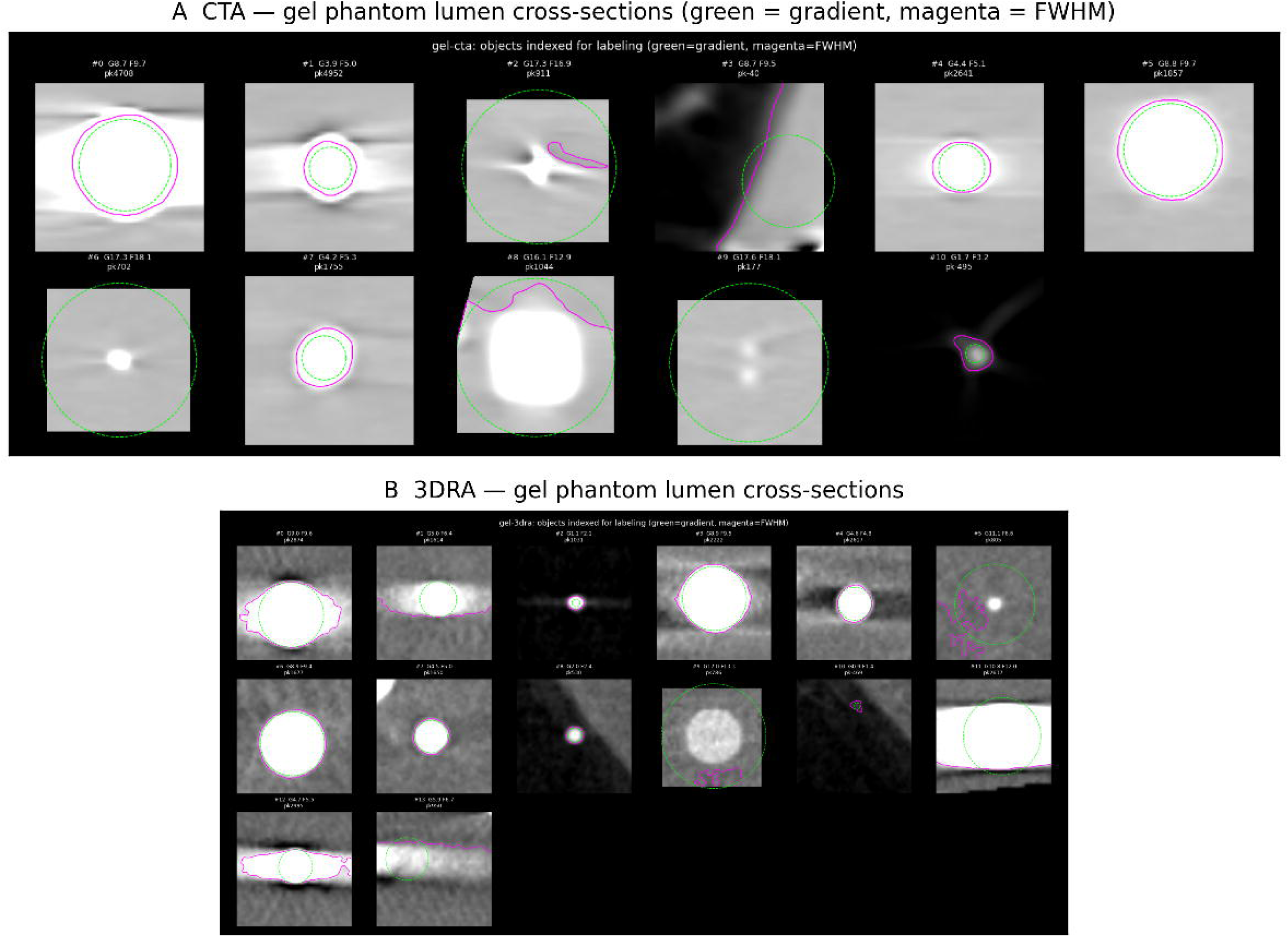
Representative gel-phantom lumen cross-sections from the blinded refinement review, on CTA (A) and 3DRA (B); each lumen shows the maximum-gradient (green) and FWHM (magenta) contour. The two criteria diverge most at the largest, most-opacified lumens, where FWHM over-reads.

Accuracy was assessed by measuring error as defined by difference between measured imaging dimensions compared to physical caliper measurements. Because each lumen contributed ∼20 nested cross-sections, a linear mixed-effects model was fitted to the cross-section-level error with a random intercept per lumen (REML); fixed-effect estimates are reported with 95% confidence intervals. Per-lumen median error, mean bias, mean absolute error (MAE), and between-lumen SD were reported descriptively, stratified by criterion, modality, surround, concentration, and size. To bound best-achievable accuracy, an operator subsequently refined contours pseudo-blinded to ground truth only calling upon the ground truth measurement when needed to adjust against gross distortion or measurement resulting from automated detection.

## RESULTS

Physical caliper sampling a uniform lumen at ∼20 levels gave near-identical diameters (within-lumen SD ∼0.04–0.05 mm for the gradient edge), indicating that essentially all measurement error is systematic rather than noise. Repeatability and accuracy by criterion are summarized in Table 1. Within-lumen SD was ≤0.24 mm for caliper-acquired physical measurements.

**TABLE 1.** Repeatability and accuracy of vessel-diameter measurement by lumen-edge criterion, versus caliper ground truth.

| Lumen-edge criterion | Repeatability (within-lumen SD, mm) | CTA bias (mm) | 3DRA bias (mm) | 2D DSA bias (mm) |
| --- | --- | --- | --- | --- |
| Maximum-gradient / manual edge† | 0.05 | +0.60 | +0.40 | +0.18 |
| FWHM | 0.05 | +1.20 | +0.98 | — |
| Fixed-HU threshold‡ | 0.05 (CTA) / 0.41 (3DRA) | +0.9 to +1.7 | -2.1 (SD 3.2); unusable | — |

*Repeatability = mean within-lumen SD of measured diameter across ~20 cross-sections of uniform phantom-1 lumens; near-zero values indicate error is systematic, not noise. Accuracy = mean signed error (measured – caliper), pooled across phantoms. †2D DSA diameter was read by a single manual edge (analogous to the gradient edge); FWHM and fixed-HU thresholds were applied only to the cross-sectional CTA/3DRA data. ‡Fixed thresholds shown at 300 HU (CTA) and 3000 (3DRA); CTA bias rose from +0.9 to +1.7 mm with dilution, and on 3DRA the variable auto-HU scale makes any absolute threshold meaningless.*
*Repeatability = mean within-lumen SD of measured diameter across ~20 cross-sections of uniform phantom-1 lumens; near-zero values indicate error is systematic, not noise. Accuracy = mean signed error (measured – caliper), pooled across phantoms. \*Fixed thresholds shown at*
300 HU (CTA) and 3000 (3DRA); CTA bias rose from +0.9 to +1.7 mm with dilution, and on 3DRA the variable auto-HU reconstruction scale makes any absolute threshold meaningless.

With the gradient edge, both CTA and 3DRA modalities measured true inner diameter to within ∼0.5 mm and erred in the same direction with over-measurement of diameters. 3DRA was modestly closer to truth and more precise than CTA with section-level gradient bias +0.40 mm on 3DRA vs +0.60 mm on CTA; MAE was 0.50 vs 0.71 mm, respectively. However, the pooled model detected no significant modality difference, and cross-vendor data showed the same trend. The observed bias was consistently directional—always an over measurement—and was minimized in the 2–5 mm diameter range (Table 2). Measurement error grew toward the extremes. Partial-volume widening was noted at the ∼1-mm resolution minimum and with the most dilute concentrations of contrast. CT saturation blooming occurred in the largest objects and most-opacified lumens with highest contrast concentrations. These findings are represented in Fig 1, Tables 1 and 2.

**TABLE 2.** Per-modality accuracy versus caliper truth. All three modalities are accurate in the mid clinical band; they diverge at the resolution floor (CTA/3DRA over-read by partial volume, 2D DSA under-reads the thin projected column) and all over-read the largest caliber.

| Modality | Measurement | n | Bias (mm) | MAE (mm) | Bias ~1.2 mm | Bias 3–4.5 mm | Bias ~8.5 mm |
| --- | --- | --- | --- | --- | --- | --- | --- |
| CTA | automatic gradient (cross-section) | 742 | +0.60 | 0.71 | +1.32 | +0.09 | +0.92 |
| 3DRA | automatic gradient (cross-section) | 940 | +0.40 | 0.50 | +0.57 | +0.11 | +0.57 |
| 2D DSA | manual edge (projection) | 145 | +0.18 | 0.46 | −0.45 | +0.10 | +0.67 |
| 2D DSA—Frontal | manual edge (projection) | 115 | +0.29 | 0.48 | −0.40 | +0.19 | +0.77 |
| 2D DSA—Lateral | manual edge (projection) | 30 | −0.21 | 0.41 | −0.60 | −0.21 | +0.17 |

*CTA/3DRA = automatic maximum-gradient edge over every valid cross-section (pooled v1+v2);*
*2D DSA = manual edge widths on objects non-overlapping in projection. Not strictly like-for-like, but each modality's error is structured and size-dependent with a shared near-zero mid band; 2D DSA additionally carries a ~0.6 mm frontal-vs-lateral magnification spread not captured by a single bias. 2D DSA lateral measurements not obtained with air phantom due to overlap from horizontal orientation.*

In a mixed-effects model pooling both phantoms (1682 cross-sections, 73 lumens), the gradient edge had an overall bias of +0.53 mm (95% CI, +0.35 to +0.72); FWHM bias was +1.08 mm (95% CI, +0.90 to +1.26), Fig 7, Table 1). No significant modality difference was detected.

Fixed-HU thresholds over-measure and were concentration-biased on CTA (bias rising from +0.9 to +1.7 mm across dilutions) and failed on 3DRA, where the variable auto-HU scale made any absolute threshold meaningless (Fig 9).

The gradient edge tracked caliper-measured physical ground truth across the full 1.2–8.5-mm range (Fig 5, Fig 6), including the sub-1.5-mm group. Residual over-measurement was concentrated at the largest caliber and minimized at sizes ≤4.5-mm. Most accurate performance was noted at 3-4.5 mm lumens, where over-measurement was 0.9-1.1mm across the three modalities. CT saturation at maximal contrast inflated over-measurement error. The descriptive surround- and size-dependent effects were not separable from phantom or scanner traits in the pooled model and are reported descriptively in Tables 1 and 2.

**FIG 5.**
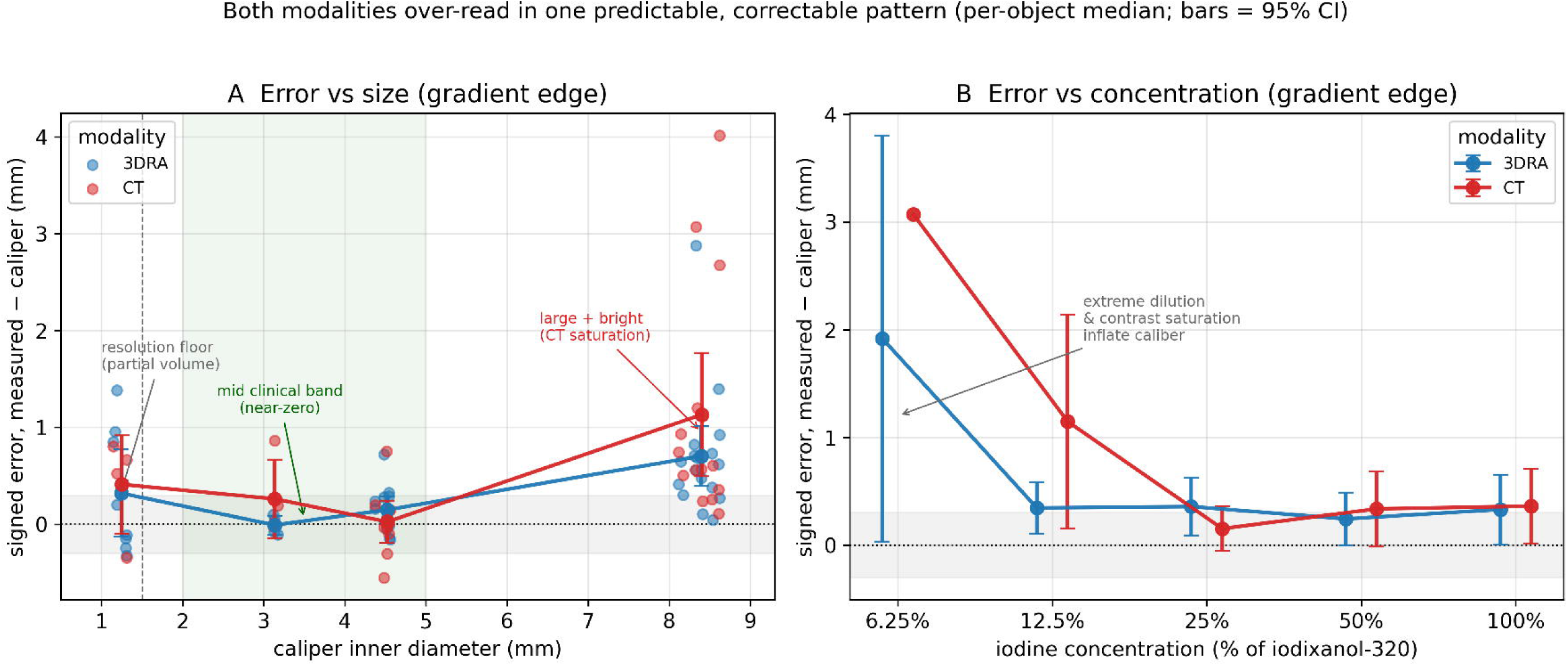
The calibration pattern. Signed gradient error (measured − caliper) versus object size (A) and versus contrast concentration (B), per modality (per-object median ± 95% CI). Both CTA and 3DRA over-read in one structured, correctable pattern: near-zero in the 2–5 mm clinical band, rising at the ∼1 mm resolution floor and at the largest, most-opacified lumens, and at extreme dilution.

**FIG 6.**
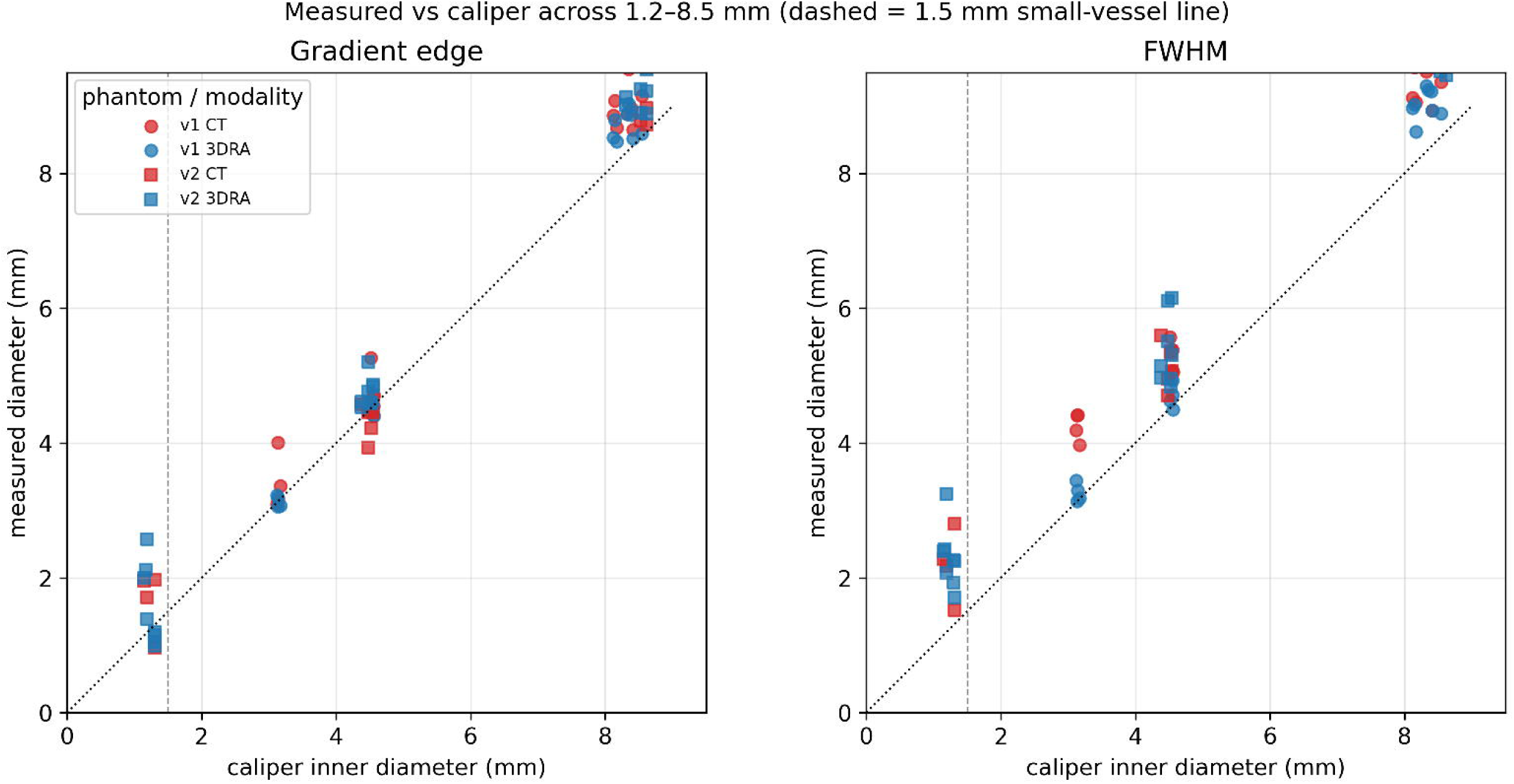
Measured versus caliper inner diameter across 1.2–8.5 mm for the gradient edge (left) and FWHM (right), both phantoms and modalities; dotted line = identity, dashed line = 1.5 mm. The gradient edge tracks identity; FWHM over-reads, most at the largest caliber.

**FIG 7.**
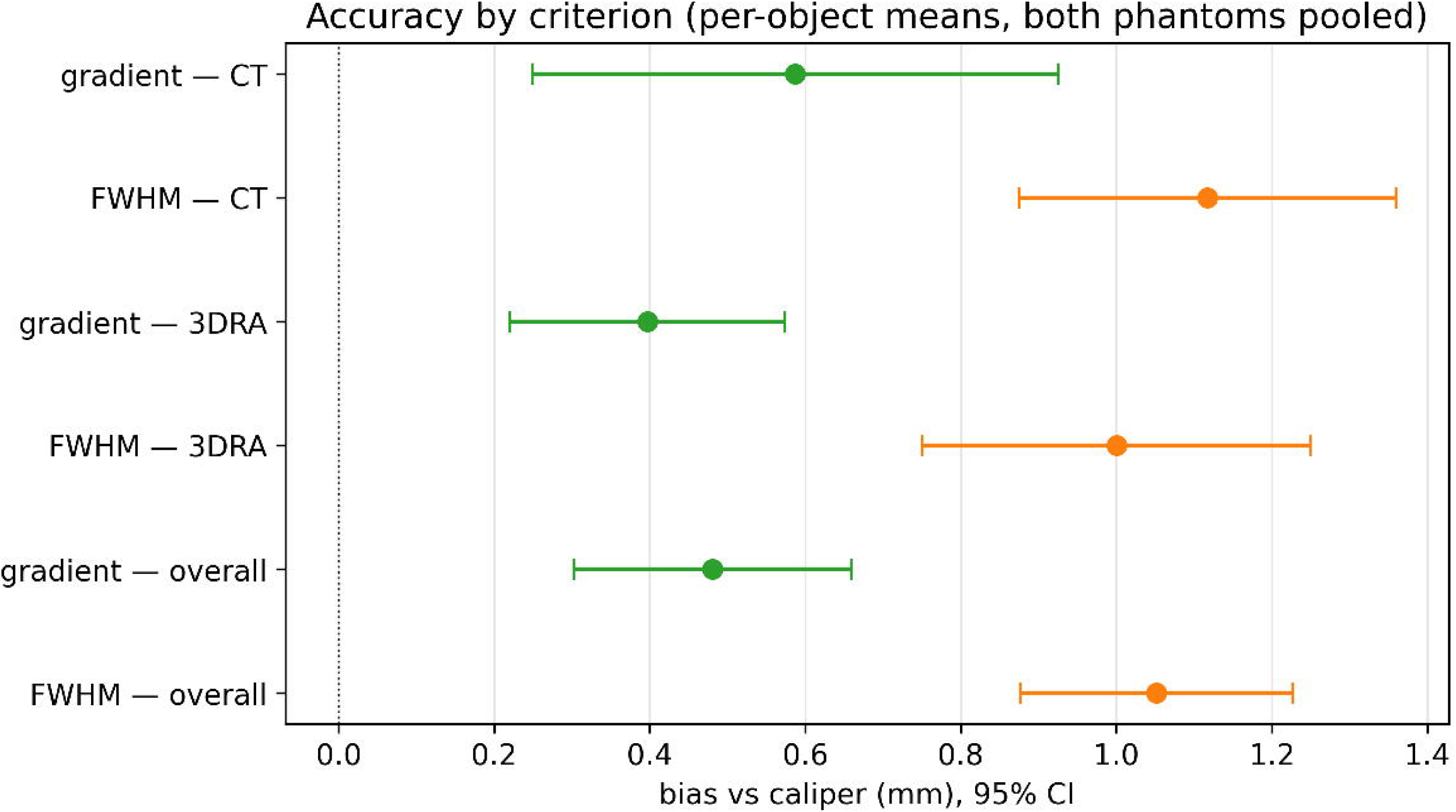
Bias versus caliper by criterion (gradient versus FWHM; overall and per modality) with 95% confidence intervals, both phantoms pooled. Non-overlapping intervals indicate the gradient edge is significantly more accurate.

Phantom 1 (GE CT + Siemens 3DRA, 16 lumens), against its own corrected cut-edge caliper, independently reproduced every phantom-2 finding on a different CT manufacturer (gradient bias +0.47 mm CT, +0.18 mm 3DRA; FWHM +1.01 and +0.43 mm). Expert refinement (pseudo-blinded predominantly to the image with ground truth hidden, with brief reference only on severely streak-distorted high-concentration 3DRA sections) reduced the gradient bias toward zero in both phantoms (e.g., phantom 1 CT +0.47→+0.17 mm; phantom-2 tubing +0.32→−0.03 mm; Medallion +1.24→+0.46 mm), indicating that the residual automatic over-measurement is largely recoverable by expert correction (Fig 8). Because of the partial unblinding, refined values bound achievable accuracy rather than independently validating it; the automatic results remain the primary, operator-independent estimate.

**FIG 8.**
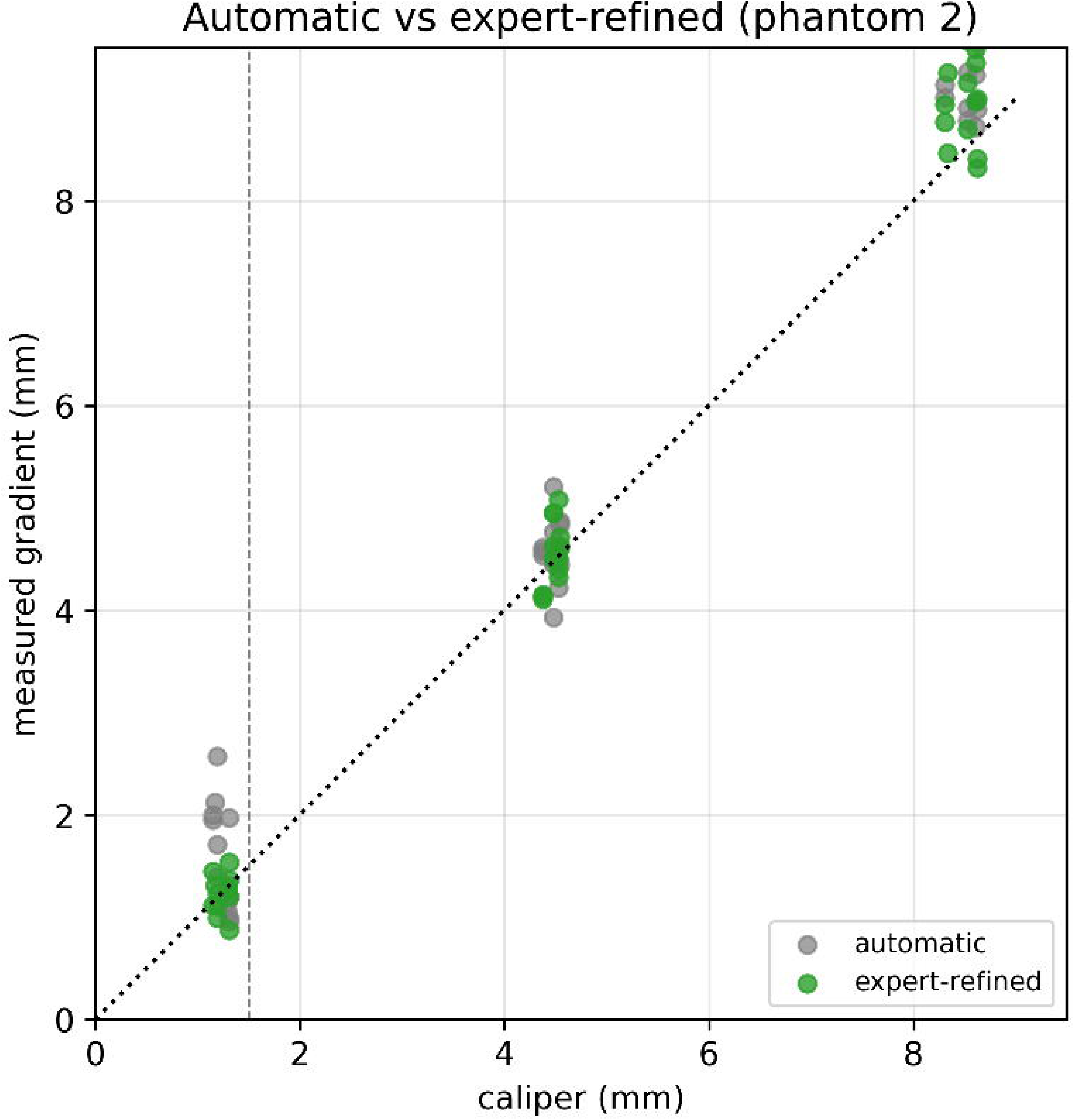
Automatic versus pseudo-blinded expert-refined gradient diameter versus caliper (phantom 2). Refinement reduces the residual over-read.

**FIG 9.**
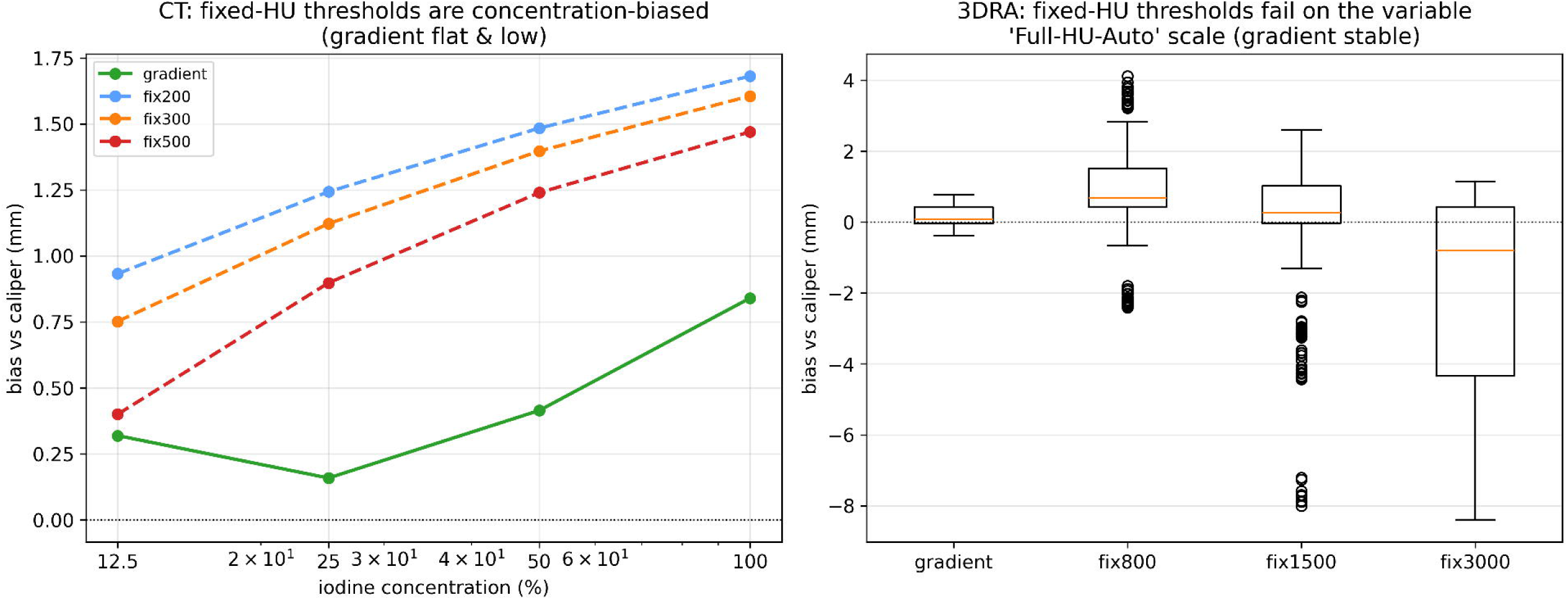
Failure of fixed-Hounsfield thresholds. Left, CT bias rises with contrast concentration for fixed thresholds while the gradient edge stays flat and low. Right, on 3DRA the fixed thresholds show catastrophic spread on the variable auto-HU scale while the gradient edge remains near zero.

Conventional 2D DSA—the modality most prior work treats as the reference standard—was itself biased against caliper-measured ground truth. Across 145 manually measured widths the overall bias was +0.18 mm (MAE 0.46), modestly smaller than the automatic cross-sectional biases but with two projection-specific failure modes (Tables 1 and 2). Notably, error occurred differently between frontal and lateral projections (Table 2), with minimal error achieved on composite 2D DSA measurements incorporating data from both projections. First, the same mid-range accuracy (+0.10 mm at 3-5 mm lumens) and large-caliber over-measurement (+0.67 mm at ∼8.5 mm lumens) as CTA/3DRA. But findings were inverted at the floor of lumen sizes. 2D DSA measurements among the sub-1.5-mm tubing lumens were under-measured by −0.45 mm. Second, a magnification signature was noted. Frontal and lateral projections of the same gel-phantom objects disagreed systematically (frontal +0.62 mm larger than lateral, scaling from +0.1 mm at the small diameter tubing to ∼+1.2 mm at 8.5 mm), because each object sits at a different distance from isocenter, and the isocenter-plane calibration cannot correct an unknown object depth. These 2D DSA findings are summarized in Fig 10.

**FIG 10.**
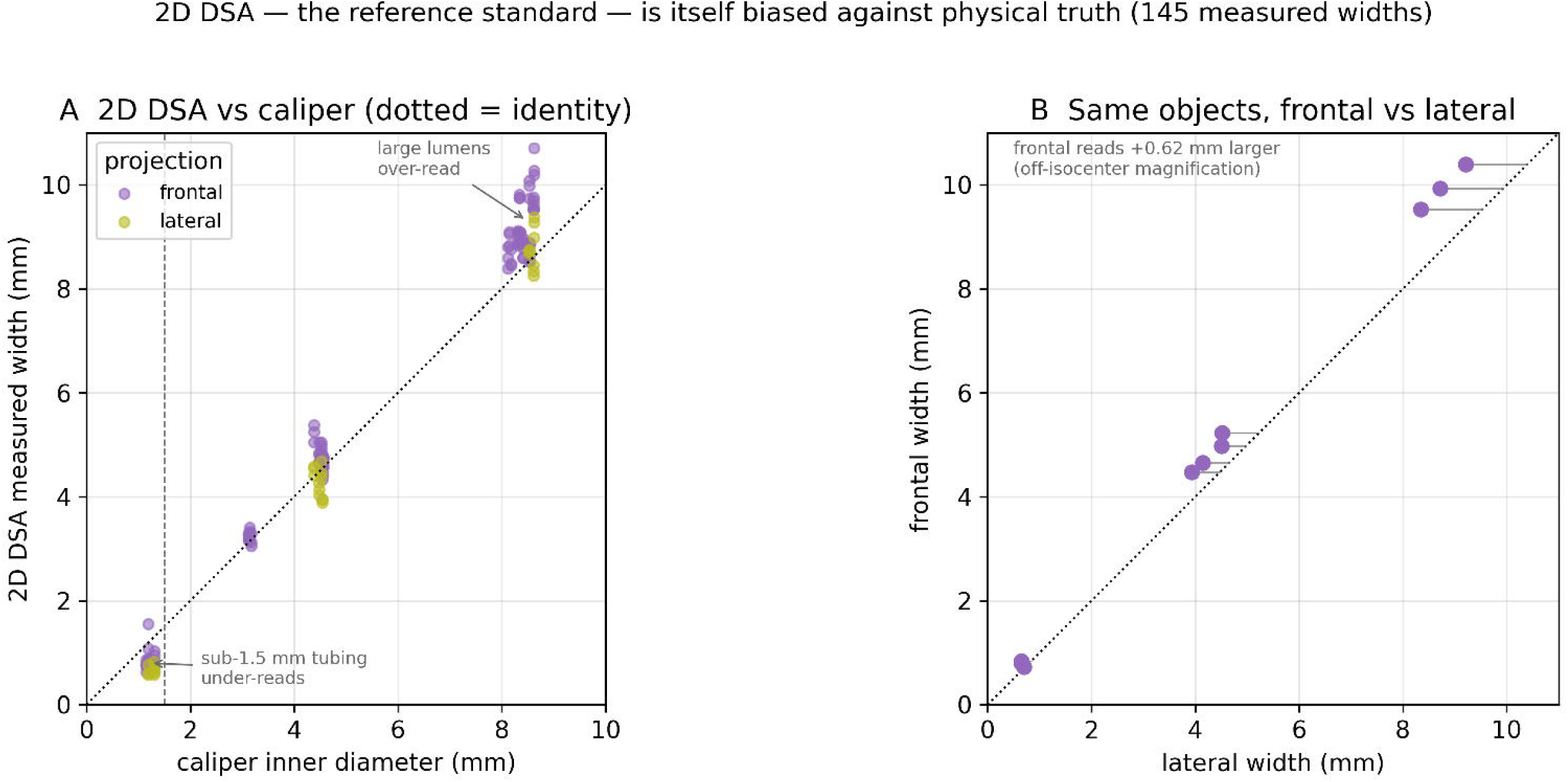
2D DSA — the reference standard — against caliper truth. A, measured versus caliper width by projection view (large-lumen over-read, sub-1.5 mm under-read; dotted = identity). B, the same gel-phantom objects measured frontally versus laterally — frontal reads +0.62 mm larger, the off-isocenter magnification signature. Together these show the reference standard is itself biased against physical truth.

## DISCUSSION

Compared against physical truth rather than another imaging modality, measurements on CTA and 3DRA both approach true intracranial-scale caliber to within ∼0.5 mm and were inaccurate in a predictable manner. The error was directional, always an over-measurement, and least pronounced in the 2–5-mm range that is most relevant clinically for measurement of large and medium intracranial arteries. Inaccuracy was amplified at extremes of both lumen size and contrast concentration. 3DRA was modestly closer to truth and more accurate than CTA, but the difference was not statistically significant. The lumen-edge criterion governed the magnitude of the bias, with maximum-gradient edge over-measured by about half as much as FWHM; gradient was also more concentration-stable compared to FWHM. Meanwhile, fixed-HU thresholding, which has been employed in some previously published non-commercial automated pipelines^18,19^, was concentration-biased on CTA and unusable on 3DRA.

Because device selection in cerebrovascular interventions hinges on ratios of the size of devices to those of vessels^1^, a sub-millimeter measurement bias might change the optimal device selection. These results support replacing fixed-HU contouring with a relative/gradient edge in automated sizing, most importantly on 3DRA, where fixed thresholds are meaningless on the auto-HU scale, and avoiding maximal contrast opacification on CT, where saturation inflates caliber, especially for larger vessels. Accuracy of the gradient edge down to ∼1.2 mm supports application to the small intracranial vessels at the lower range of those routinely accessed and intervened upon. However, adaptation of measurement regimes to reflect these results should be undertaken sensibly. Current sizing recommendations for most devices, when based on imaging data, likely reflect bias in imaging reconstructions demonstrated herein. These recommendations likely warrant calibration against known measurements that account for such imaging biases. For instance, an optimized catheter-to-vessel ratio for mechanical thrombectomy based on over-estimated arterial lumens on CTA obtained during triage may need to be revisited considering these findings. Refinement through future studies including phantom data is certainly warranted, particularly if such work includes measurements of non-deformable interventional devices to compare against their true physical dimensions.

The value of measuring against physical truth is that it converts an unknown error into a known, correctable one. Because the bias is systematic in direction and patterned by size and concentration rather than stochastic, ground rules for interpretation of automated measurements can begin to be constructed. Automated measurements may be most trustworthy in the ∼2–5 mm luminal diameter range, requiring only mild downward correction, while such correction must be increased in both the largest, brightest vessels, where saturation inflates caliber, and in small lumens near the ∼1-mm resolution floor and in extreme dilution situations. This calibration, rather than verdicts on which modality or criteria wins outright, is most appropriate for the development of automated image processing platforms.

FWHM is widely treated as the partial-volume-correct diameter criterion of choice^11^, but that status rests on idealizing assumptions, including a symmetric point-spread function, a clean step edge, and a stable, known peak-to-background amplitude. Under real contrast variation those assumptions fail. The 50%-of-peak crossing drifts as luminal intensity changes, so FWHM’s over-measurements are both larger and more concentration-sensitive, carrying roughly twice the bias seen with gradient edge. This persisted across modalities and surrounds in the phantoms interrogated. The maximum-gradient edge keys on the radius of steepest transition, independent of absolute amplitude and background, leading to more accurate measurements with more robust performance across contrast concentrations and, in particular, for small, high-contrast lumens such as those seen in medium and small intracranial vessels. Prior FWHM endorsements were validated in simulation or against angiographic references^20^. This analysis, to the authors’ knowledge, represents the first head-to-head test against physical truth across contrast concentration and the small-vessel range, and it favors the gradient edge.

The clinical literature reports that CTA underestimates large vessels in tissue relative to DSA^9,10^, whereas this analysis shows air phantom DSA, 3DRA, and CTA images over-measured relative to caliper-based ground truth measurements. This finding reflects the surround (air versus tissue) and the reference (DSA versus physical caliper). The finding is refined by measuring 2D DSA against the same caliper truth. This typically employed reference standard was itself biased. 2D DSA measurement bias was +0.18 mm overall, over-measuring large lumens, under-reading sub-1.5-mm tubing, and reading ∼0.6 mm larger frontally than laterally on the same object due to magnification artifacts. Previous literature that benchmarked CTA against DSA thus compared two biased modalities. Methodological refinement is added in this study with the comparison to a physical standard, whose images and their measurements are closest to ground truth. All three modalities land within ∼0.5 mm but by different, separately-correctable mechanisms—partial volume and saturation for cross-sectional CTA/3DRA, magnification and projection artifacts for 2D DSA. Over-measurement occurred for most modalities, contrast concentrations, and lumen sizes, with the exception being small lumens on 2D DSA, i.e. the tubing cohort with ∼1-1.5mm diameter. This likely occurs because a thin, faint contrast column in projection is edged tightly with edge detection that is difficult to discern. This study represents, to the authors’ knowledge, the first calibration of CTA, 3DRA, and 2D DSA against caliper-based measurements of known phantom structures across contrast concentration and in the sub-2-mm range.

At maximal 3DRA 100% contrast, streak/starburst artifact rendered some cross-sections ovoid; no edge criterion can recover a true circle from a geometrically corrupted image. Such sections are best flagged by a circularity gate or human review and excluded rather than measured in such considerations. Consistent with this, expert refinement recovered most of the residual large-caliber over-read, arguing that an automated platform should retain expert qualitative review options. This is particularly relevant for power-injection of contrast for obtaining 3DRA, during which most or all of the intra-luminal blood is temporarily replaced by injected contrast. This is in distinction to 2D DSA injections during which smaller total volumes of contrast are typically injected, either by power injection or hand injection. Higher concentrations of contrast are likely to lead to such artifact that may lead to spurious measurements that could in turn result in suboptimal procedural results due to potential changes in device selection.

The current study has several limitations. Lumen numbers per cell were modest (6–16), limited by the ability to confine multiple devices within a confined space sufficiently close to isocenter to obtain optimal images. Limiting close positioning to minimize artifact affecting adjacent structures and leakage from some devices during acquisition limited the usable lumen numbers. A trend toward improved measurement accuracy for CT when moving from air to a gel/saline surround warrants further analysis and could potentially achieve significance with a larger sample size. This would be relevant for design and execution of future phantom studies. The expert-refined analysis was pseudo-blinded and bounds best-achievable rather than independently validated results, which may introduce bias. However, complete blinding would have excluded multiple datasets that were screened out by automatic methods due to artifacts. Rigid plastic walls differ from thin arterial walls; however, the gradient edge keys on the contrast-to-wall interface and is therefore relatively insensitive to the surrounding material beyond it. Furthermore, other factors that can affect vessel caliber in vivo, such as pulsatility, motion, vessel-wall composition, and distensibility are not modeled by the present technique.

Additionally, while efforts were made to avoid stretching or compression of the extension tubing used for the smallest lumens, the nonrigid material could have been over-measured by calipers due to this limitation in materials. Additionally, the flexibility of tubing precluded maintaining a straight configuration in the phantom and limited ability to reliably measure a cross section at some segments of tubing.

## CONCLUSIONS

Against physical ground truth, CTA, 3DRA, and 2D DSA all measure intracranial-scale vessel caliber to within ∼0.5 mm, but each departs from measured ground truth in a consistent, correctable manner. Maximum-gradient lumen edge minimizes measurement biases and is more concentration-insensitive than FWHM, with the latter previously considered the conventional preference. Fixed-Hounsfield contour tracing performed poorly throughout all circumstances and should be avoided, most notably on 3DRA. 2D DSA errors resulted from geometric magnification artifacts and under-measurement at smaller lumen sizes below 1.5-mm. Previous acceptance of 2D DSA as the reference standard should be recalibrated based on these findings. Characterizing measurement inaccuracy of each modality suggests the consistent error modes are likely correctable, and automated measurement schemes could be calibrated to provide more accurate dimensions. Further investigation is warranted.

## Data Availability

All data produced in the present study are available upon reasonable request to the authors

## ABBREVIATIONS

CTA: CT angiography
3DRA: 3D rotational angiography
DSA: digital subtraction angiography
FWHM: full-width at half maximum
HU: Hounsfield unit
CI: confidence interval
MAE: mean absolute error

